# Linkage of National Congenital Heart Disease Audit data to hospital, critical care and mortality national datasets to enable research focused on quality improvement

**DOI:** 10.1101/2021.11.07.21265266

**Authors:** Ferran Espuny Pujol, Christina Pagel, Katherine L. Brown, James C. Doidge, Richard G. Feltbower, Rodney C. Franklin, Arturo Gonzalez-Izquierdo, Doug W. Gould, Lee J. Norman, John Stickley, Julie A. Taylor, Sonya Crowe

**Author notes:** **Corresponding Author** Ferran Espuny Pujol, Clinical Operational Research Unit, Department of Mathematics, University College London, London, UK, 4 Taviton Street, London WC1H 0BT, UK.

## Abstract

**Objectives:** To link five national datasets (three registries, two administrative) and create longitudinal health care trajectories for patients with congenital heart disease (CHD), describing the quality and the summary statistics of the linked dataset.

**Design:** Bespoke linkage of record-level patient identifiers across five national datasets. Generation of spells of care defined as periods of time-overlapping events across the datasets.

**Setting:** National congenital heart disease audit (NCHDA) procedures in public (NHS) hospitals in England and Wales, paediatric and adult intensive care datasets (PICANet and ICNARC-CMP), administrative hospital episodes (HES inpatient, outpatient, A&E), and mortality registry data.

**Participants:** Patients with any CHD procedure recorded in NCHDA between April 2000 and March 2017 from public hospitals.

**Primary and secondary outcome measures:** Primary outcomes: Number of linked records, number of unique patients and number of generated spells of care (e.g. inpatient stays, outpatient appointments).

Secondary outcomes: Quality and completeness of linkage.

**Results:** There were 143,862 records in NCHDA relating to 96,041 unique patients. We identified 65,797 linked PICANet patient admissions, 4,664 linked ICNARC-CMP admissions, and over 6 million linked HES episodes of health care (1.1M Inpatient, 4.7M Outpatient). The 96,041 unique patients had 4,908,153 spells of care comprising 6,481,600 records after quality checks. Considering only years where datasets overlapped, 95.6% surgical procedure records were linked to a corresponding HES record, 93.9% paediatric (cardiac) surgery procedure records were linked to a corresponding PICANet admission, and 76.8% adult surgery procedure records were linked to a corresponding ICNARC-CMP record.

**Conclusions:** We successfully linked four national datasets to the core dataset of all CHD procedures performed between 2000 and 2017. This will enable a much richer analysis of longitudinal patient journeys and outcomes. We hope that our detailed description of the linkage process will be useful to others looking to link national datasets to address important research priorities.

**Strengths and limitations of this study:** *Strengths:*

- We linked five national established, high quality, datasets using bespoke methods for the pre-processing of identifiers and establishing matches to maximise linkage
- In our final dataset, data consistency has been checked at patient level using year and month of birth, postcodes, and diagnosis codes, and also clinically sense-checked at spell level for spells containing congenital heart procedures.
- We created meaningful spells of care for each patient in the dataset covering inpatient and outpatient interactions with secondary and tertiary care, covering up to 20 years of life of patients with congenital heart disease (CHD), representing an important step to understanding patient care for people with CHD.

*Limitations:*

- Data completeness, quality and availability were worse in earlier years, meaning that linkage was poorer for earlier eras.
- We do not yet have data on hospital care for patients outside England or on longer term adult follow-up for patients whose full CHD history is captured, since most cardiac procedures start in early live – the national CHD audit (NCHDA) started on April 2000.

## Introduction

Measuring, reporting and learning from patient outcomes should drive quality improvement (QI), but this is particularly challenging for lifelong conditions where outcomes need to be interpreted in the context of different phases of treatment, changing treatment options, changing service provision and the natural evolution of disease.[1,2] Given the complex longitudinal care trajectories of such patients, rich datasets and careful multi-disciplinary analysis are required to understand how patients interact with health services and to identify relevant outcomes and meaningful variations. This then provide opportunities for more targeted QI. Services for congenital heart disease (CHD) provide one such example. They span a patient’s lifetime, but their quality in the UK is mainly measured by 30-day survival following children’s heart surgery or catheter-based procedures. This is no longer a sufficient proxy and a more sophisticated approach is required.[3]

Information on patients with CHD, and their utilisation of specialized care services in England and Wales, is not available in a single dataset. Since April 2000, the main source of information on the early outcomes of therapeutic paediatric and congenital cardiovascular procedures for CHD patients in UK has been the *National Congenital Heart Disease Audit* (NCHDA).[4,5] Submission is mandatory for all centres and data quality is subject to external validation. The key feature of this dataset is the detailed recording of cardiac-related diagnosis and procedural information using the *European Paediatric and Congenital Cardiac Code* (EPCCC) short list descriptors.[6]

By linking NCHDA with other national datasets, both validated registries and administrative, we aimed to build a unique combined dataset for understanding patient journeys through the secondary and tertiary healthcare system. The four relevant national datasets are: the paediatric intensive care audit network (PICANet) for patient admissions to paediatric intensive care units;[7] the case mix programme from the intensive care national audit & research centre (ICNARC-CMP) for patient admissions to adult intensive care units;[8] death registrations from the Office for National Statistics (ONS); and hospital episode statistics (HES) routine administrative data on admitted patient care (APC), accident and emergency (A&E) attendances, and outpatient (OP) appointments at NHS hospitals in England.[9,10]

The research project LAUNCHES QI: “Linking Audit and National datasets in Congenital Heart Services for Quality Improvement” aims to: describe patient trajectories through secondary and tertiary care; identify useful metrics for driving QI and informing commissioning and policy; explore variation across services to identify priorities for QI. In this paper, our objective is to describe the methods used to link the NCHDA data to HES, ONS, PICANet and ICNARC-CMP datasets, and report the general characteristics, strengths and limitations of the resulting LAUNCHES dataset. The process and challenges involved in the application for the approvals needed to link the LAUNCHES datasets has been described elsewhere.[11]

## Methods

### Data

The core dataset in LAUNCHES is **NCHDA**,[4,5] from which we obtained data for all records between 1 April 2000 and 31 March 2017 (Figure 1). Each record relates to a single CHD procedure carried out in public hospitals in England and Wales. Most patients are resident in England and Wales, but patients from Northern Ireland and Scotland and overseas are also represented. NCHDA provides detailed demographic, diagnosis and procedural information for CHD procedures in children and adults, as well as short-term survival outcomes (in-hospital and at 30 days).[12] Table S1 in Supplementary Material contains all NCHDA fields that we obtained for LAUNCHES.

**Figure 1.**
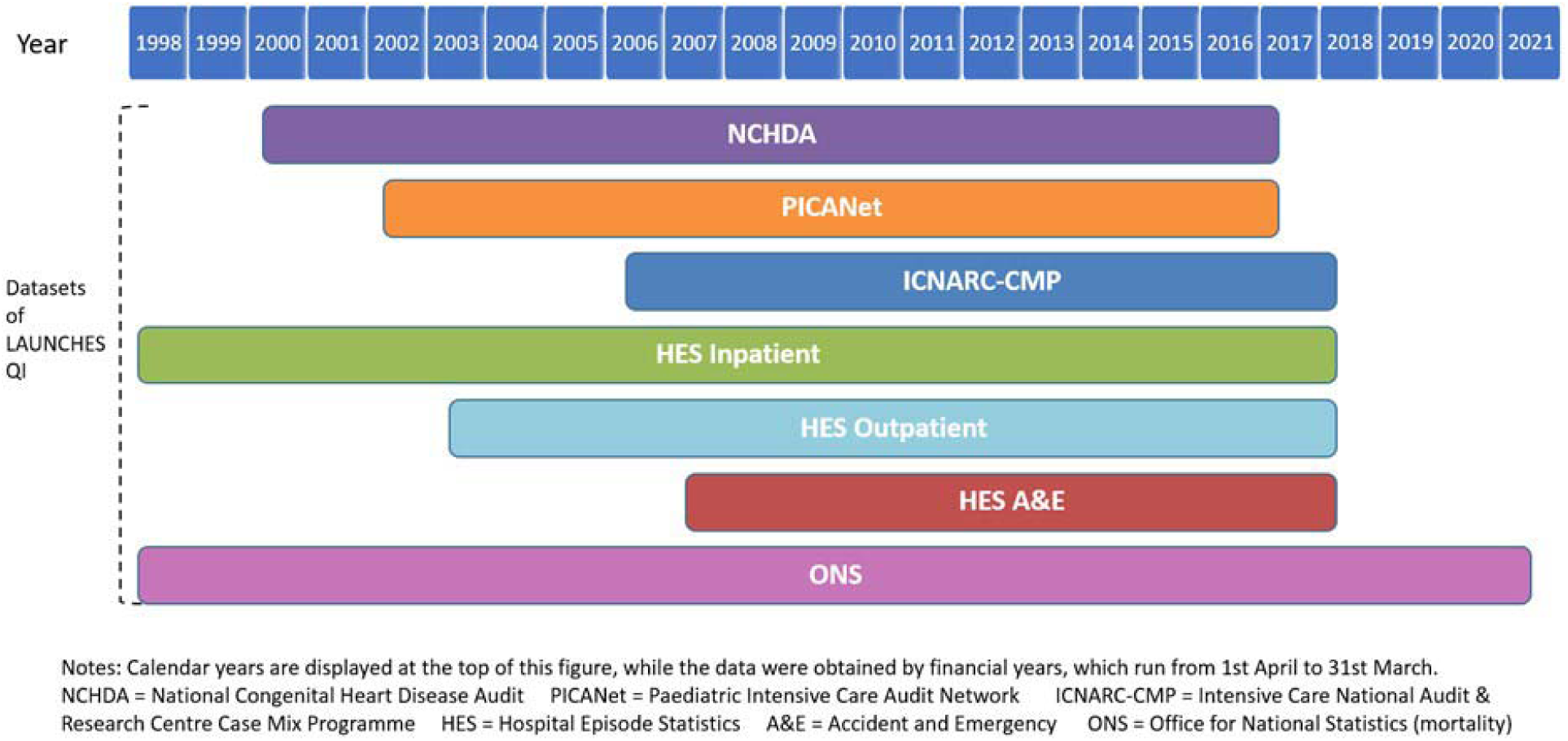
Datasets and years covered to make up the LAUNCHES dataset.

We applied to link to the following *Hospital Episode Statistics* **(HES)** datasets (Figure 1): Admission Patient Care (APC) inpatient (not limited to cardiac) admissions to hospitals in England from financial years 1998/99 (starting 1 April 1998) and 2017/18 (ending 31 March 2018); HES Outpatient (OP) appointments between financial years 2003/04 (first year available) and 2017/18; HES Accident and Emergency (AE) attendances between financial years 2007/08 (first year available) and 2017/18.[9,10,13] Tables S2, S3, and S4 contain all HES fields that we obtained for LAUNCHES.

*The Office for National Statistics* (**ONS**) mortality data is the most complete source for the assessment of patient survival, recording all deaths registered in England and Wales.[14] Linked to HES data,[15] we obtained the ONS life status of patients of patients resident in England and Wales. See Table S5 for all ONS fields.

The *Paediatric Intensive Care Audit Network* (**PICANet**) contains records for all children admitted to paediatric intensive care units within UK and Republic of Ireland.[7] We requested all PICANet admissions in England up to March 2017 that could be linked to records in NCHDA (see Table S6 for all PICANet fields).

The *Case Mix Programme* (**CMP**) collects data from adult general critical care units in England, Wales and Northern Ireland.[8,16] We requested all ICNARC-CMP admissions up to August 2018 that could be linked to records in NCHDA (see Table S7 for all ICNARC-CMP fields).

The selected HES years correspond to all years of HES data with available HES identifiers (HES IDs) and NHS numbers (see HES Data Dictionary [17]) at the application time, where HES APC year 1997/98 was not requested because we were informed that NHS numbers were largely missing (55.5%).

No dates of patient events were requested, other than year and month of birth (Tables S1 to S6). Instead, ages (in years) to 4 decimal places at each event were requested from each data provider to facilitate construction of detailed health care trajectories (enabling ordering of multiple events on the same day) while minimising identifiability of the linked data.

### Data identifiers used for linkage

Table 1 lists the identifiers used for linkage, the datasets each were present in, and any pre-linkage processing that was undertaken. NHS numbers have some limitations,[18,19] particularly that they are likely to be missing for overseas patients or those from Scotland and Northern Ireland. Hospital identifiers are unique to a patient, and records with the same hospital identifier will relate to the same patient. But hospital identifiers change between hospitals and so are not useful for linking patient records across different hospitals. In the absence of a matching NHS number or hospital patient identifier, we used date of birth, name and postcode to identify records as pertaining to the same patient but only if all three matched across records. We categorised the quality of each identifier for each record as: valid (for linkage), invalid, or missing (Table 1).

**Table 1.** Identifiers used for linkage.

| Identifier | Description and processing undertaken | Dataset |
| --- | --- | --- |
| NHS number | NHS numbers are 10-digit identifiers assigned to people registered for NHS care in England, Wales, or the Isle of Man. They are assigned to patients soon after birth (since year 2002) or the first time they receive NHS care or treatment.[30]<br>*Processing:* removed non-numeric characters and blanks.<br>*Invalid values:* 10-digit numbers that are all the same; dummy value “2333455667”; format “n00000000n” (e.g. “6000000006” ).[15]<br>*Valid values:* Not invalid (above) and satisfying the checksum digit check.[31] | NCHDA,<br>PICANet,<br>ICNARC-CMP,<br>HES/ONS |
| Hospital Patient ID | Hospitals use their own local patient identifiers, which in combination with the centre ID constitute a unique patient identifier that we refer to as “hospital patient identifier”. A patient can have multiple hospital identifiers across their records e.g. associated with care in different hospitals at different times.<br>*Processing:* standardised the centre ID values, and removed blanks, leading zeroes and leading/trailing special characters from the local patient identifiers.[15]<br>*Valid values:* any value was considered valid. | NCHDA,<br>PICANet |
| Date of birth (DoB) | Date of birth of the patient is available as recorded in the datasets<br>*Processing:* standardised the format to day/month/year (e.g. 17/11/2007”).<br>*Invalid values:* Any date after 01/04/2017 or before 01/01/1895. Not equal to either 01/01/1901 or 31/12/1899.[15]<br>*Valid values:* Not invalid (see above) and a feasible date. | NCHDA,<br>PICANet,<br>ICNARC-CMP,<br>HES/ONS |
| Name/surname | *Processing:* converted to upper case; removed prefixes and titles (e.g. MISS, MSTR, MASTER, MRS, MS, MR, MAST, DR, SGT, SHEIKHA, SULTANA, SHEIKH, SULTAN), removed generic values (e.g. BABY, INFANT, TWIN, TRIPLETS, BOY, GIRL, NAME1, NAME2 etc.) Removed special characters (apostrophes and accents).<br>*Valid values:* non-empty values (after processing the fields). | NCHDA,<br>PICANet |
| Postcode | *Processing:* converted to uppercase, removed blanks and special characters (only alphanumeric characters allowed).<br>*Valid values:* postcodes included in the historical list of postcodes from the Organisation Data Service [32] and not corresponding to country postcodes (starting with “ZZ”) and not from an NHS trust site.[33] | NCHDA,<br>PICANet,<br>ICNARC-CMP,<br>HES/ONS |

### Linkage method

We developed an algorithm to link NCHDA data both internally (to identify records pertaining to the same person within NCHDA), and externally, to records in the other datasets. Our hierarchical method, shown in Figure 2, treated NHS number and Hospital Patient ID as primary identifiers, whilst date of birth, patient name and postcode were treated as weaker identifiers. The possible linkage states when comparing a processed identifier across two records were:

**Figure 2.**
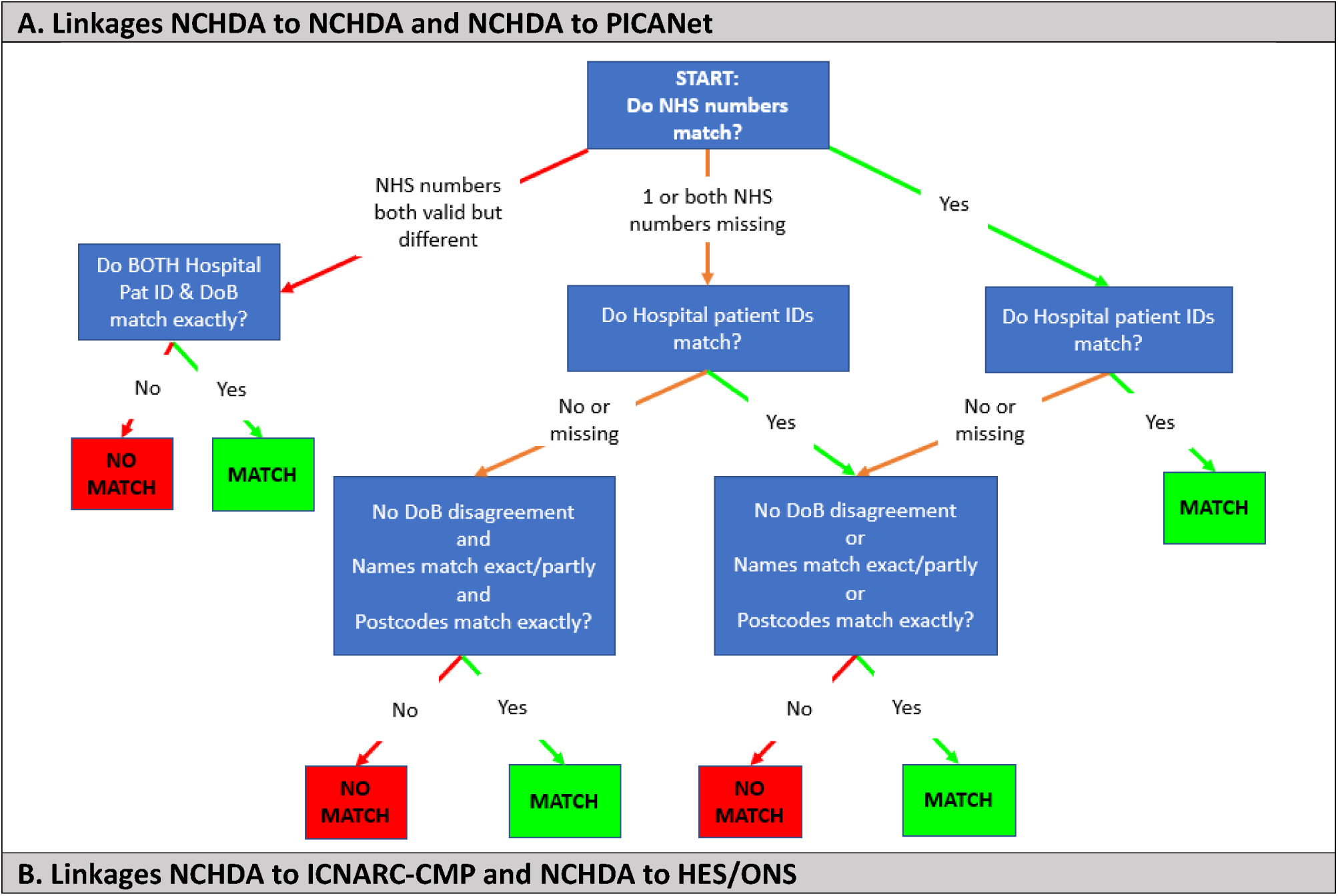
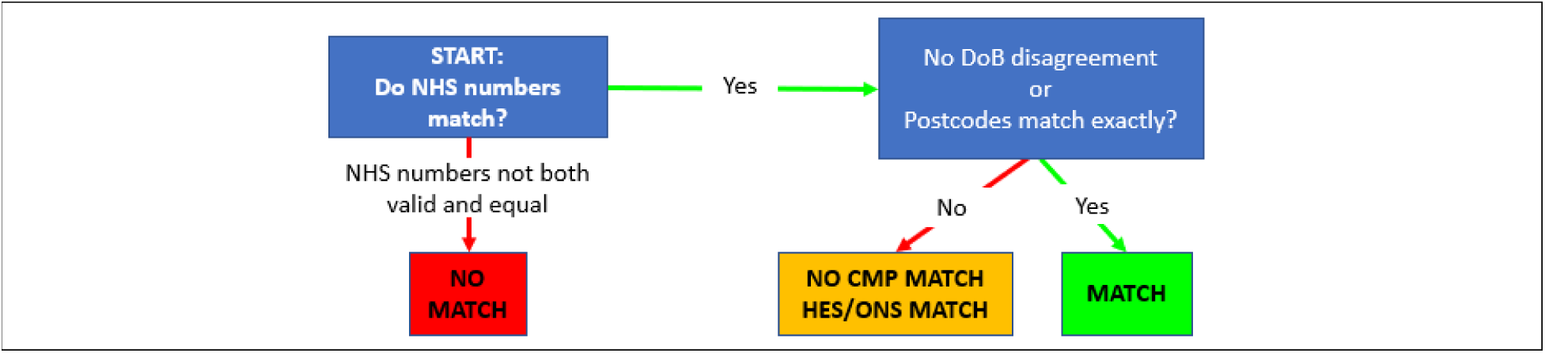
The linkage algorithm for deciding whether two records pertain to the same patient. Note: “No DoB disagreement” means that the dates of birth either match (exactly or partially) or one or both of those dates are missing.

- Exact agreement, if each identifier was valid and they were exactly the same.
- Partial agreement, only used for valid dates of birth and names, and defined in detail below.
- Any missing, if either or both identifiers were missing or invalid.
- Disagreement, if both values were valid and non-missing but did not match (exactly or partially).

Two valid dates of birth (DoB) were considered to be in partial agreement if either: the two DoB values were no more than 5 days apart; the two DoB values were not the same, but either two components (i.e. YYYY, MM or DD) of the two DoB values matched or two components of the two DoB values matched when the MM and DD parts of one of them were swapped. Partial agreement of names occurred between two records if there were previous and current versions of names and at least one matched the other record.

An auxiliary lookup table (Table S8) between NCHDA organisations and Paediatric Intensive Care Units (PICUs) was used by PICANet when comparing hospital patient identifiers as part of the NCHDA to PICANet linkage (Figure 2A), given that the two datasets use different names for centres.

For NCHDA to ICNARC-CMP linkage, two records were matched by ICNARC only if there was exact agreement of NHS numbers and either the DoB did not disagree or postcodes matched exactly (Figure 2B). NCHDA to HES/ONS linkage was performed by NHS Digital and required the exact match of NHS numbers (agreement in postcode was reported but not required). See Table S9 for the HES/ONS linkage method.

Finally, note that all linkages were done at record level. This resulted in many-to-many record matches that were resolved to identify records as pertaining to the same patient across all five datasets once pseudonymised datasets had been received at UCL.

### Data flows

Record-level patient identifiers in the core dataset (NCHDA) were sent for linkage via secure transfer to each of the three data controllers for the other four datasets, along with a study-specific pseudonymised record identifier. Each data controller then searched for records within their datasets with matching patient identifiers, and returned the pseudonymised, clinical data (without patient identifiers) for all records that had at least one match to an NCHDA record to University College of London (UCL) Clinical Operational Research Unit (CORU). We used secure transfer and all data are stored in the UCL data safe haven, which complies with the NHS Information Governance Toolkit. Only pseudonymised study-specific record and patient IDs were shared with or stored at UCL. Linkage results were provided as lists of corresponding pairs of records with a code indicating the quality of linkage for each record-to-record match (concatenated agreement category for each identifier).

### Patient level consistency and quality assurance

The national audit body (NICOR) identified unique patients within the NHCDA using the linkage algorithm, and then checked for inconsistencies on site as part of data quality assurance. Inconsistencies in dates of birth (missing values, procedures before birth, different dates of birth for a same patient) were identified and sent to submitting hospitals for correction and were then revised by NICOR. Cleaned record identifiers were then sent for linkage to the other data processors. An additional internal detailed clinical review was undertaken of pairs of records that were not linked but similar to some extent (e.g. those pairs solely agreeing in NHS number) and pairs of records linked but with only moderate agreement in identifiers (e.g. pairs with matched names, dates of birth and postcode but NHS numbers missing), and internal patient categorisation updated.

Both HES and PICANet have their own internal unique patient IDs across records. Pseudonymised versions of these were included in the returned records. We then assessed the level of agreement between the identified patients from the NCHDA and patient identifiers from the linked PICANet and HES datasets. PICANet and HES patients linked to more than one LAUNCHES patient were discussed with each processor and patient categorisation was revised on a case-by-case basis.

### Spells of care and completeness of linkage

Once the linked dataset was created, we combined overlapping events into ‘spells of care’. Gaps of less than 24 hours were considered to be overlapping, since times of events were not routinely collected and so records could have a 12-hour uncertainty in either direction. Figure 3 illustrates an example of event records that would be combined into a single (paediatric) spell.

**Figure 3.**
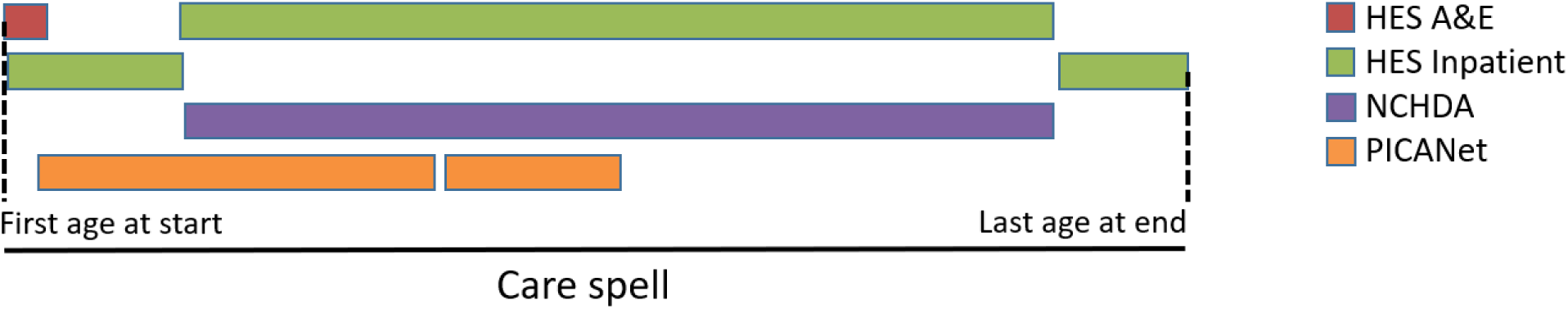
Example of Care spell consisting of several time-overlapping events involving different services.

Cardiac surgeries typically require intensive care recovery. Catheter-based interventions and diagnostic procedures are far less likely to require ICU admission. Our first consistency check was to look at how many spells containing a cardiac surgery procedure also contained an accompanying ICU stay, enabling an assessment of the completeness of linkages from NCHDA to PICANet and NCHDA to ICNARC-CMP. While we would not expect 100% of NCHDA surgeries to have a linked record, we would expect a high proportion to. A second consistency check was for HES linkage completeness. We would expect a HES linked record (either inpatient admission or outpatient attendance) to be part of the same spell as any NCHDA procedure, as long as the NHCDA record had a valid NHS number. In addition, at least one of the ICD-10 diagnostic codes used within HES for inpatient admissions should denote CHD for HES records linked to NCHDA surgical procedures (a list of valid congenital codes and other cardiac non-congenital codes that are sometimes used for CHD patients is provided in Table S10).

### Ethical approval

LAUNCHES received ethical approval from the Health Research Authority (reference: IRAS 246796) and the Confidentiality Advisory Group (reference: 18/CAG/0180).

### Patient and public involvement statement

We have patient and public representatives on the independent study advisory group. The advisory group were consulted on linkage design and execution and approved the process.

## Results

### Quality of identifiers in each dataset

The NCHDA dataset contained 143,862 CHD records of which 94.7% had valid NHS numbers. Unsurprisingly, the percentage of valid NHS numbers was higher for patients with residence in England (98.8%) or Wales (99.1%) as determined by their postcode at the time of procedure. The breakdown of NHS numbers by residence is given in Table S11. PICANet records for patients born before 14 October 2001 were available only if they had a PICANet event between 14 October 2014 and 13 October 2019, due to the terms of the PICANet Health Research Authority (HRA) Confidentiality Advisory Group (CAG) approval for processing identifiable information.[20] There were 179,791 PICANet records available for linkage, of which 90.5% had valid NHS numbers. Hospital patient Identifiers were available for 100% of NCHDA and PICANet records, as were dates of birth; names/surnames were available for 99.6% and 98.9% of records, respectively, and postcodes were valid for 95.% and 97.2% of records. ICNARC-CMP had 1,853.568 records of which 88.7% had valid NHS numbers. The total of records and percentage of valid NHS numbers for HES data were: 314,445,082 (93.8%) for HES Inpatient, 1,288,711,692 (98.0%) for HES Outpatient, and 194,572,279 (93.3%) for HES A&E. We did not know the quality of identifiers in ONS mortality data, which we obtained linked to HES data. The quality of the identifiers improved over time (Table S12).

### Linked datasets before quality assurance

There were 6,408,673 records across the final component datasets before any quality assurance was carried out (Table S13 in the Appendix), with each non-NCHDA record linked to at least one NCHDA record.

#### Quality of the record level linkage

The use of a bespoke method for linking NCHDA-NCHDA and NCHDA-PICANet records (Figure 2A) allowed us to identify more linked records than had we relied solely on NHS numbers:

- 95.0% of the NCHDA-NCHDA matches and 92.3% of the NCHDA-PICANet matches were identified by an exact agreement of NHS numbers;
- 4.9% of the NCHDA-NCHDA and 7.0% of the NCHDA-PICANet matches were identified by exact agreement in hospital patient identifiers (allowing for missing NHS number);
- 0.1% of the NCHDA-NCHDA and 0.7% of the NCHDA-PICANet matches were identified by other options of our bespoke linkage algorithm.

### Patient-level results

There were 47,753 internal NCHDA linked records (out of a total of 143,862 NCHDA records), representing patients with more than one recorded procedure within the NCHDA dataset.

Once patients had been defined across NCHDA records, 649 inconsistencies in dates of birth affecting 219 patients were detected and corrected. There was a very high level of agreement between the identified patients from the linked PICANet data and the LAUNCHES linkage definition of patients: only 7 PICANet patients (0.0% of the 34,507 linked PICANet patients) were linked to two LAUNCHES patients each. Investigation of those cases by each audit resulted in a further minor revision. In a similar exercise, we excluded 88 HES IDs (0.1% of the total 89,098 linked HES IDs) that were linked to two LAUNCHES patient IDs each. It was not possible to determine which HES records corresponded to each patient (mainly because they pertained to twins). Inconsistencies between 42 HES and NCHDA patients linked with disagreement in year-month of birth and postcode were also resolved.

This detailed review of linked NCHDA records resulted in a final total of 96,041 unique patients with a total of 6,381,600 records (Table 2). 66,453 patients (69.2%) had at least one NHCDA record as children (age at procedure under 16). 29,588 patients (30.8%) had all their NHCDA records as adults. 90,678 patients (94.5%) were linked to at least one external dataset: 91.5% of patients had some form of HES/ONS record, 35.9% had at least one linked PICANet record, and 3.6% had at least one linked ICNARC-CMP record. 5,363 patients (5.6% of all NCHDA patients) were not linked to any other dataset, mainly due to: missing NHS number; residence not recorded or outside England; and/or record from before 2003 when data quality was poorer. The final linked dataset covers up to 20 years of life of patients, with a median (IQR) coverage of 12 (6,16) years for 88,275 patients with no known age of death, and 4 (1, 12) years for 7,766 patients with known age of death.

**Table 2.** Number of linked records in each dataset after quality assurance, by estimated financial year

| **Financial Year** | **NCHDA** | **PICANet** | **ICNARC-CMP** | **HES Inpatient** | **HES Outpatient** | **HES A&E** | **Total** |
| --- | --- | --- | --- | --- | --- | --- | --- |
| **1998** | 0 | 0 | 0 | 16,431 | 0 | 0 | 16,431 |
| **1999** | 0 | 0 | 0 | 19,811 | 0 | 0 | 19,811 |
| **2000** | 6,421 | 15 | 2 | 29,113 | 0 | 0 | 35,551 |
| **2001** | 6,161 | 11 | 1 | 33,210 | 0 | 0 | 39,383 |
| **2002** | 6,137 | 952 | 0 | 36,870 | 0 | 0 | 43,959 |
| **2003** | 7,402 | 3,226 | 0 | 42,805 | 132,364 | 0 | 185,797 |
| **2004** | 6,968 | 3,464 | 0 | 45,314 | 149,544 | 0 | 205,290 |
| **2005** | 7,684 | 3,828 | 0 | 50,097 | 176,383 | 0 | 237,992 |
| **2006** | 8,152 | 4,052 | 6 | 52,001 | 195,655 | 0 | 259,866 |
| **2007** | 7,984 | 4,136 | 154 | 56,577 | 223,402 | 23,268 | 315,521 |
| **2008** | 8,294 | 4,275 | 215 | 59,782 | 254,476 | 27,482 | 354,524 |
| **2009** | 8,719 | 4,748 | 273 | 65,190 | 292,972 | 32,732 | 404,634 |
| **2010** | 8,987 | 4,891 | 388 | 69,084 | 322,196 | 35,862 | 441,408 |
| **2011** | 9,102 | 5,103 | 407 | 70,564 | 347,096 | 38,854 | 471,126 |
| **2012** | 9,013 | 5,176 | 411 | 70,908 | 368,160 | 41,598 | 495,266 |
| **2013** | 9,593 | 5,435 | 473 | 71,781 | 406,805 | 42,830 | 536,917 |
| **2014** | 9,639 | 5,435 | 447 | 72,751 | 440,554 | 44,913 | 573,739 |
| **2015** | 11,492 | 5,546 | 629 | 75,959 | 468,434 | 47,219 | 609,279 |
| **2016** | 12,114 | 5,504 | 686 | 72,899 | 476,727 | 46,885 | 614,815 |
| **2017** | 0 | 0 | 572 | 51,814 | 424,473 | 43,432 | 520,291 |
| **All years** | **143,862** | **65,797** | **4,664** | **1,062,961** | **4,679,241** | **425,075** | **6,381,600** |

Note: Financial years (running from April to March) were estimated using the ages at events and the estimated date of birth (we took day 15<sup>th</sup> of the known month of birth as date of birth). The estimation is likely wrong for 27 records from PICA Net and ICNARC-CMP with estimated year pre-2002, but we could not fix the needed ages or dates of birth at the time of submission (such inconsistencies are likely to be excluded in future analyses).

### Spell-level results

We identified 4,908,153 spells of care for the 96,041 patients in the LAUNCHES dataset. Only 2.6% of the spells contained one or more NCHDA procedures compared to 99.7% of spells which included at least one HES record. 1.0% of spells included at least one PICANet record, and 0.1% of spells included at least one ICNARC-CMP record. Patients had a median (IQR) of 3.4 (1.8, 6.3) spells per year, with a median (IQR) of 0.1 (0.1,0.3) spells with NCHDA procedures per year. This high level of health care interaction was expected in this population, since patients with CHD require regular specialist follow-up.

### Sense checking the completeness of the linkage

#### PICANet

93.9% (42,512/45,265) of paediatric cardiac surgeries, and 11.2% (2,047/18,268) of paediatric catheter-based procedures, were linked to an associated PICANet record where linkage was in principle feasible. The high proportion of cardiac surgeries linked provides reassurance that we achieved high quality linkage between the two datasets (since we would not expect 100% of surgeries to require PICU care). The proportion of catheter-based procedures linked matched clinical experience since a minority of catheter-based interventions require pre- or postprocedural intensive care.

#### ICNARC-CMP

76.8% (906/1,180) of adult cardiac surgeries (and 2.6% (69/2,610) of catheters) were linked to ICNARC-CMP when the procedures were post-March 2009 at centres submitting regularly to ICNARC, and where a valid NHS number was recorded. Unfortunately, many hospitals carrying out congenital heart procedures submitted very few records to ICNARC-CMP over the time period of this study. This means that for all cardiac surgeries where ICNARC-CMP data would have been available (post 2009 with a valid NHS number), only 16.5% (1,193/7,234) were linked to an associated CMP record.

#### HES

95.6% (122,278/127,932) NHCDA procedure records (either surgical or catheter) with a valid NHS number and performed in an English public hospital were linked to an associated HES record, mostly Inpatient records. ONS age at death was provided for 7,228 patients. In a total of 53,769 spells which included both NCHDA surgical procedures and an associated HES Inpatient record, 94.6% of HES records had CHD ICD-10 diagnostic codes from Table S10, 3.8% had only acquired heart diagnoses (plausible miscoding of CHD), and 1.6% had other diagnostic codes.

These consistency checks provide assurance that, where linkage was theoretically possible, we achieved excellent linkage.

## Discussion

### Principal findings

We have described a bespoke linkage algorithm, alongside quality, completeness and consistency checks, that we used to identify 96,041 unique patients across 143,862 NCHDA cardiac procedure records and to link their records to 65,797 PICU admissions, 4,664 adult Intensive Care admissions and 6,167,277 HES (inpatient, outpatient and A&E) records.

While most of the linked records were identified using matching NHS numbers, a significant proportion (around 5%) were identified using other identifiers, highlighting the value of using additional identifiers. Close collaboration with each audit and NHS Digital meant that we could further check the quality of the linkage and further refine the identification of unique patients across records, improving the overall quality of the linked dataset.

The quality of recorded identifiers used for linkage improved markedly over time as did the quality of resulting linkage. 90,678 (94.5%) patients had records that were linked to at least one other dataset. We identified 4,908,153 spells of care for the 96,041 patients. The final linked dataset (6,381,600 records) covers up to 20 years of life of patients, with a median (IQR) coverage of 12 (6,16) years for 88,275 patients with no known age of death, and 4 (1, 12) years for 7,766 patients with known age of death.

Patients had a median (IQR) of 3.4 (1.8, 6.3) spells of care (either an inpatient stay or an outpatient event) per year. This frequent interaction with secondary and tertiary care outside of NCHDA procedures (only 2.6% spells of care included an NCHDA procedure) highlights the necessity and value of linking specialised validated procedure-based registry records (NCHDA) to other administrative and audit datasets to understand and potentially improve services for CHD.[21,22]

### Strengths and weaknesses

All linked datasets were national established, high quality, datasets. We designed a bespoke linkage method and data processors carefully prepared the identifiers for linkage in a consistent way to maximise matching. In our final dataset, data consistency has been checked at patient level using year and month of birth, postcodes, and diagnosis codes, and also clinically sense-checked at spell level for spells containing congenital heart procedures.

Each of the datasets used for linkage were available for different years. Additionally, PICANet’s HRA CAG policy of data anonymisation restricted linkage feasibility for some patients, HES data only covered hospitals in England, and ICNARC-CMP dataset was of limited utility since many specialised adult cardiac intensive care units did not submit to ICNARC-CMP for most or all of the time period. More adult cardiac ICUs submit to ICNARC-CMP every year and so future linkage should be much more complete.

The linked dataset covers at most 20 years of life of patients. While this represents an important step to understanding patient care for people with CHD, we do not yet have data on longer term adult follow-up for patients whose full CHD history is captured (i.e. those born after 2000), since most cardiac procedures start in early life.

### Comparison with other studies

In the UK, the Infant Heart Study linked an NCHDA cohort to PICANet data to explore risk factors for poor outcomes (one year) after hospital discharge for infants undergoing heart surgery between years 2005 and 2010. [23,24] ONS mortality was included as part of NCHDA at that time, and the linkage to PICANet was carried out using just NHS number. A study looking at Differences in access to Emergency Paediatric Intensive Care and care during Transport (DEPICT) linked together PICANet, ICNARC-CMP, and HES/ONS. NHS numbers were the primary identifiers used for matching.[25–27] Our bespoke linkage algorithm improved the approach based on NHS numbers, with 7.7% of the total NCHDA-PICANet matches obtained using agreement in other identifiers.

### Implications for clinicians and policy-makers

The NHCDA database is highly specialized and procedure based. The linked intensive care and hospital datasets provide a much wider and more complete picture of the interactions CHD patients have with secondary and tertiary care throughout their lives. In particular, the outpatient data means loss to follow up in transition from child to adult services and/or during adulthood can be explored. The linked data of validated registries with administrative databases will facilitate the identification of appropriate outcomes for reporting and routine monitoring CHD services at all ages, including resource utilisation, and to develop methods of QI that take into account differences in risk across case-mix.[28]

### Unanswered questions and future research

The NCHDA data set only contains information for CHD patients that have at least one procedure. This means that when considering overall health service journeys of people living with CHD, we miss those who never have a procedure (either because disease is considered too mild or because it is too severe for correction). The ongoing CHAMPION project will use the National Congenital Anomaly and Rare Disease Registration Service (NCARDRS) dataset to estimate the number of children born with CHD, or that have an antenatal diagnosis but do not survive pregnancy (termination or in-utero death).[28,29] In future, linkage to NCARDRS might allow assessment of outcomes and health care journeys for the complete patient cohort.

## Conclusion

We successfully linked five national datasets to achieve a large, high quality combined dataset spanning 20 years that will allow rich exploration of the health care journeys of patients with congenital heart disease. We hope that this detailed description will be useful to others looking to link national datasets to address important research priorities.

## Data Availability

This paper describes the linkage of five national datasets and does not present results based on
analysis of that data. The linked data is held and processed in the Data Safe Haven under strict
governance requirements and signed data sharing agreements. It cannot be shared with others
without significant amendments to ethics, CAG and data sharing agreements.

## Acknowledgments

We would like to thank the data application teams at PICANet, ICNARC, NICOR, HQIP, and NHS Digital for their help and guidance as we negotiated the data application system.

## Funding statement

This study is supported by the Health Foundation, an independent charity committed to bringing about better health and health care for people in the UK (Award number 685009).

## Competing interests statement

None declared.

## Author contributions statement

All authors contributed to the design of the bespoke linkage algorithm. R code was developed by FEP for the processing, quality assessment and linkage of NCHDA records (R code will be posted at github). Audit collaborators at PICANet (LN) and ICNARC (JD) adapted the code to perform the linkage. All authors contributed to quality and consistency assurance of the linkage and dataset. The clinical sense-checking of linked records and spells of care was performed by RF and KB. CP and SC conceived of and led the study. FEP wrote the first draft of the manuscript and all authors edited, commented and approved the final draft.

## Data sharing statement

This paper describes the linkage of five national datasets and does not present results based on analysis of that data. The linked data is held and processed in the Data Safe Haven under strict governance requirements and signed data sharing agreements. It cannot be shared with others without significant amendments to ethics, CAG and data sharing agreements.

## Notes

### Competing Interest Statement

The authors have declared no competing interest.

## References

1 Rogers L, Brown KL, Franklin RC, et al. Improving Risk Adjustment for Mortality After Pediatric Cardiac Surgery: The UK PRAiS2 Model. The Annals of Thoracic Surgery 2017;104:211–9. doi:10.1016/j.athoracsur.2016.12.014

2 Rogers L, Pagel C, Sullivan ID, et al. Interventions and Outcomes in Children With Hypoplastic Left Heart Syndrome Born in England and Wales Between 2000 and 2015 Based on the National Congenital Heart Disease Audit. Circulation 2017;136:1765–7. doi:10.1161/CIRCULATIONAHA.117.028784

3 New Congenital Heart Disease Review: Final Report. NHS England 2015. https://www.england.nhs.uk/wp-content/uploads/2015/07/Item-4-CHD-Report.pdf

4 NICOR | Congenital Heart Disease in Children and Adults (Congenital audit). https://www.nicor.org.uk/national-cardiac-audit-programme/congenital-heart-disease-in-children-and-adults-congenital-audit/ (accessed 23 Aug 2021).

5 Franklin R, Wang J, Ajayi S, et al. National Congenital Heart Disease Audit. 2020 Summary Report (2018/19 Data). Healthcare Quality Improvement Programme (HQIP) 2020.

6 Franklin RCG, Anderson RH, Daniëls O, et al. Report of the Coding Committee of the Association for European Paediatric Cardiology. Cardiol Young 2002;12:1–8. doi:10.1017/S1047951100012208

7 Universities of Leeds & Leicester. PICANet – Paediatric Intensive Care Audit Network for the UK and Ireland. https://www.picanet.org.uk/ (accessed 23 Aug 2021).

8 Harrison DA, Brady AR, Rowan K. Case mix, outcome and length of stay for admissions to adult, general critical care units in England, Wales and Northern Ireland: the Intensive Care National Audit & Research Centre Case Mix Programme Database. Critical Care 2004;9:S1. doi:10.1186/cc3745

9 Herbert A, Wijlaars L, Zylbersztejn A, et al. Data Resource Profile: Hospital Episode Statistics Admitted Patient Care (HES APC). Int J Epidemiol 2017;46:1093–1093i. doi:10.1093/ije/dyx015

10 Boyd A, Cornish R, Johnson L, et al. Understanding Hospital Episode Statistics (HES). London, Uk: : CLOSER 2018. https://www.closer.ac.uk/wp-content/uploads/CLOSER-resource-understanding-hospital-episode-statistics-2018.pdf

11 Taylor JA, Crowe, S., Espuny Pujol, F., et al. The road to hell is paved with good intentions: the experience of applying for national data for linkage and suggestions for improvement. BMJ Open 2021;11:e047575. doi:10.1136/bmjopen-2020-047575

12 White O, Stickley J. National Congenital Heart Disease Audit. Data Manual For dataset version 6.1 – March 2020 Revision. 2020.

13 Hospital Episode Statistics (HES). NHS Digital. https://digital.nhs.uk/data-and-information/data-tools-and-services/data-services/hospital-episode-statistics (accessed 23 Aug 2021).

14 Linked HES-ONS mortality data. NHS Digital. https://digital.nhs.uk/data-and-information/data-tools-and-services/data-services/linked-hes-ons-mortality-data (accessed 23 Aug 2021).

15 Health and Social Care Information Centre, HSCIC. A Guide to Linked Mortality Data from Hospital Episode Statistics and the Office for National Statistics. Health and Social Care Information Centre, HSCIC 2015.

16 ICNARC – About the CMP. https://www.icnarc.org/Our-Audit/Audits/Cmp/About (accessed 23 Aug 2021).

17 Hospital Episode Statistics Data Dictionary. NHS Digital. https://digital.nhs.uk/data-and-information/data-tools-and-services/data-services/hospital-episode-statistics/hospital-episode-statistics-data-dictionary (accessed 23 Aug 2021).

18 Moser K, Hilder L. Assessing quality of NHS Numbers for Babies data and providing gestational age statistics. Health statistics quarterly 2008.

19 Adoption and gender re-assignment processes - Primary Care Support England. https://pcse.england.nhs.uk/help/registrations/adoption-and-gender-re-assignment-processes/ (accessed 14 Apr 2021).

20 PICANet. Policy of data anonymisation. https://www.picanet.org.uk/wp-content/uploads/sites/25/2019/11/PICANet-ongoing-data-anonymisation.pdf (accessed 30 Jun 2021).

21 Pasquali SK, Peterson ED, Jacobs JP, et al. Differential Case Ascertainment in Clinical Registry vs. Administrative Data and Impact on Outcomes Assessment in Pediatric Heart Surgery. Ann Thorac Surg 2013;95:197–203. doi:10.1016/j.athoracsur.2012.08.074

22 Jacobs JP, Franklin RCG, Béland MJ, et al. Nomenclature for Pediatric and Congenital Cardiac Care: Unification of Clinical and Administrative Nomenclature – The 2021 International Paediatric and Congenital Cardiac Code (IPCCC) and the Eleventh Revision of the International Classification of Diseases (ICD-11). Cardiology in the Young 2021;31:1057–188. doi:10.1017/S104795112100281X

23 Crowe S, Ridout DA, Knowles R, et al. Death and Emergency Readmission of Infants Discharged After Interventions for Congenital Heart Disease: A National Study of 7643 Infants to Inform Service Improvement. Journal of the American Heart Association;5:e003369. doi:10.1161/JAHA.116.003369

24 Brown KL, Wray J, Knowles RL, et al. Infant deaths in the UK community following successful cardiac surgery: building the evidence base for optimal surveillance, a mixed-methods study. Southampton (UK): : NIHR Journals Library 2016. http://www.ncbi.nlm.nih.gov/books/NBK363028/ (accessed 23 Aug 2021).

25 Ramnarayan P, Evans R, Draper ES, et al. Differences in access to Emergency Paediatric Intensive Care and care during Transport (DEPICT): study protocol for a mixed methods study. BMJ Open 2019;9:e028000. doi:10.1136/bmjopen-2018-028000

26 Seaton SE, Ramnarayan P, Davies P, et al. Does time taken by paediatric critical care transport teams to reach the bedside of critically ill children affect survival? A retrospective cohort study from England and Wales. BMC Pediatrics 2020;20:301. doi:10.1186/s12887-020-02195-6

27 Seaton SE, Ramnarayan P, Pagel C, et al. Impact on 30-day survival of time taken by a critical care transport team to reach the bedside of critically ill children. Intensive Care Med 2020;46:1953–5. doi:10.1007/s00134-020-06149-5

28 CHAMPION project, NIHR PR-R20-0318-23001. https://fundingawards.nihr.ac.uk/award/PR-R20-0318-23001 (accessed 23 Aug 2021).

29 National Congenital Anomaly and Rare Disease Registration Service (NCARDRS). GOV.UK. https://www.gov.uk/government/collections/national-congenital-anomaly-and-rare-disease-registration-service (accessed 23 Aug 2021).

30 What is an NHS number? nhs.uk. 2018.https://www.nhs.uk/using-the-nhs/about-the-nhs/what-is-an-nhs-number/ (accessed 14 Apr 2021).

31 NHS NUMBER. https://datadictionary.nhs.uk/attributes/nhs_number.html (accessed 14 Apr 2021).

32 Office for National Statistics data. NHS Digital. https://digital.nhs.uk/services/organisation-data-service/data-downloads/office-for-national-statistics-data (accessed 14 Apr 2021).

33 Other NHS organisations. NHS Digital. https://digital.nhs.uk/services/organisation-data-service/data-downloads/other-nhs-organisations (accessed 14 Apr 2021).

