## Supplemental Tables for "Linkage of National Congenital Heart Disease Audit data to hospital, critical care and mortality national datasets to enable research focused on quality improvement"

### Supplementary Tables

**Table S1** NCHDA fields obtained.

| NCHDA field name | Description |
|---|---|
| 1.00 DataSet Version | The version of the dataset the data was collected for and submitted to NICOR |
| 1.01 Hospital identifier | The identifier allocated to the hospital by NICOR. The software should set this field without any user involvement. |
| 1.07 Patient Gender | Identifies the genotypical sex of the patient. |
| 1.08 Patient Ethnic Group | Identifies the patient's ethnic origin. |
| 1.09 Patient Admin status | Type of admission, i.e. from UK public health service or other mode of entry to the service |
| 2.01 Diagnosis | The preprocedural diagnosis of the patient |
| 2.02 Previous Procedure | Relevant previous procedures |
| 2.03 Weight | The patients weight in kg at the time of procedure to two decimal places. |
| 2.03b Height | Height at time of procedure in cm |
| 2.04 Antenatal Diagnosis | Diagnosis detected prior to birth from prenatal scans |
| 2.05 Preprocedure seizures | Any preprocedural convulsions/seizures requiring medication |
| 2.06b Comorbidity present | A comorbidity is the presence of one or more additional disorders (or diseases) co-occurring with a primary disease or disorder; or the effect of such additional disorders or diseases. |
| 2.07 Comorbid Conditions | Identifies the specific comorbid condition |
| 2.08 Preprocedure systemic ventricular ejection fraction | Categorises the percentage of the blood emptied from the systemic ventricle at the end of the contraction. Data may have been derived from angiography, echocardiography, nuclear imaging, magnetic resonance imaging etc. Use this metric to define ventricular function in patients with functionally single ventricle anatomy. |
| 2.09 Preprocedure subpulmonary ventricular ejection fraction | Categorises the percentage of the blood emptied from the subpulmonary ventricle at the end of the contraction. Data may have been derived from angiography, echocardiography, nuclear imaging, magnetic resonance imaging etc. Do not use this metric for patients with functionally single ventricle anatomy. |
| 3.01b Procedure urgency | Categorises the patient in terms of the urgency |
| 3.01c Unplanned reoperation | Used to identify cases that aren't part of the planned pathway for that patient. |
| 3.04 First operator grade | The grade of the secondary operator or assistant. |
| 3.06 First assistant grade | The grade of the secondary operator or assistant. |
| 3.07 Type of Procedure | Defines the group the procedure should be included in. |
| 3.08 Sternotomy Sequence | Incremental count of the number of sternotomies that the patient has undergone. |
| 3.09 Operation performed | The EPCC short codes that describe the procedure |
| 3.10 Total bypass time | The total duration of cardiopulmonary bypass used during the procedure. |
| 3.11 Total bypass cross clamp time | The total duration of aortic cross clamp during the procedure. |
| 3.12 Total circulatory arrest time | The total duration of circulatory arrest during the procedure. |
| 3.13 Catheter procedure duration | The operative time taken. |
| 3.14 Total fluoroscopy time | The total time fluoroscopy was used during the procedure |
| 3.15 Total fluoroscopy dose | The total fluoroscopy dose during the procedure |

| | |
|---|---|
| 3.16 Procedure Report Or Comment | Accompanying text that can help describe the procedure in cases where coding is thought to be inadequate. |
| 4.03 Discharge status | The status of the patient at discharge from your hospital. |
| 4.04 Discharge Destination | The immediate destination following discharge from your hospital |
| 4.05 Postprocedure seizures | Any postprocedural convulsions/seizures requiring medication |
| 4.07 Duration of postoperative intubation | Duration of postoperative intubation associated with a procedure. |
| 4.08 Postoperative complications | Significant postoperative complications following surgery |
| 4.09 Attribution of death | The attribution of death to a procedure |
| 5.01 Device Manufacturer | The manufacturer of any implanted devices. |
| 5.02 Device model | The model numbers of any implanted device. |
| 5.04 Device Size | The size of any devices implanted. |
| 6.01 Preprocedure NYHA status | The patient's preprocedural NYHA status. |
| 6.02 Preprocedure smoking status | The patient's preprocedural smoking status |
| 6.03 Preprocedure diabetes | The patient's preprocedural diabetes status |
| 6.04 History of pulmonary disease | The patient's preprocedural pulmonary disease status |
| 6.06 Preprocedural ischaemic heart disease | The patient's preprocedural ischaemic heart disease status |
| 7.01 Preprocedural valve or septal defect or vessel size | The preprocedural size of the valve or septal defect or vessel size |
| 7.02 Sizing balloon used for septal defect closure Y/N | Was a sizing balloon used for septal defect occlusion |
| 7.03 Number of stents or coils | The number of stents and/or coils deployed |
| 7.04 Catheterisation complication severity rating | Classifies the severity of the most major catheter complication. |
| 7.05 Catheterisation complications | Significant postprocedural complications following a cardiac catheter |
| **LAUNCHES derived field name** | **Description** |
| LAUNCHESrecID | LAUNCHES record identifier |
| LAUNCHESpatID | LAUNCHES patient identifier |
| LAUNCHESpatID_rev | LAUNCHES patient identifier revised |
| Qcode | Record identifiers' quality code |
| dob.year | Year of birth derived from *1.06 Patient Date of Birth* |
| dob.month | Month of birth derived from *1.06 Patient Date of Birth* |
| country | Country of patient's residence derived from *1.10 Patient Post Code* |
| IMD2004.rank | IMD2004 rank of patient's residence derived from *1.10 Patient Post Code* |
| IMD2004.decile | IMD2004 decile of patient's residence derived from *1.10 Patient Post Code* |
| IMD2007.rank | IMD2007 rank of patient's residence derived from *1.10 Patient Post Code* |
| IMD2007.decile | IMD2007 decile of patient's residence derived from *1.10 Patient Post Code* |
| IMD2010.rank | IMD2010 rank of patient's residence derived from *1.10 Patient Post Code* |
| IMD2010.decile | IMD2010 decile of patient's residence derived from *1.10 Patient Post Code* |
| IMD2015.rank | IMD2015 rank of patient's residence derived from *1.10 Patient Post Code* |
| IMD2015.decile | IMD2015 decile of patient's residence derived from *1.10 Patient Post Code* |
| Raop | Age in years (4 decimal places) at procedure derived from *3.01 Date/Time procedure* |
| RFLAGtop | Record FLAG: time of procedure present? |
| Raodis | Age in years (4 decimal places) at discharge derived from *4.01 Date of Discharge* |
| Raod | Age in years (4 decimal places) at death derived from *4.02 Date of Death* |

Note: The NCHDA linked data and linkage quality report were received on 23 September 2019. The revised ages and anonymised patient key identifiers were received on 27 February 2019.

**Table S2** HES inpatient (HES APC) fields obtained.

| HES Inpatient field name | Description |
|---|---|
| ADMIFLAG | Admission episode flag |
| ADMIMETH | Method of admission |
| ADMISORC | Source of admission |
| AEKEY | Record identifier |
| BIRSTAT_N | Birth status |
| CLASSPAT | Patient classification |
| DIAG_NN | All Diagnosis codes |
| DISDEST | Destination on discharge |
| DISFLAG | Discharge episode flag |
| DOMPROC* | Trust derived dominant procedure |
| ENCRYPTED_HESID | Patient identifier - HES generated (encrypted version) |
| EPIKEY | Record identifier |
| EPIORDER | Episode order |
| ETHNOS* | Ethnic category |
| ETHRAW* | Ethnic character (audit version) |
| FIRSTREG | First regular day or night admission |
| HRG_N.N | Healthcare resource group: version 3.1 |
| HRGNHS | Trust derived HRG value |
| HRGNHSVN | Version No. of Trust derived HRG |
| IMD04 | IMD Index of Multiple Deprivation |
| IMD04_DECILE | IMD Decile Group |
| IMD04RK | IMD Overall Rank |
| MAINSPEF | Main specialty |
| OPERTN_NN* | Primary Operative Procedure Codes |
| PROCODE5 | Provider code (5 character) |
| PROCODET | Provider code of treatment |
| PROTYPE | Provider type |
| RESGOR | Government Office region of residence |
| RESGOR_ONS | Government office region of residence (ONS) |
| RURURB_IND | Rural/Urban Indicator |
| SEX* | Sex of patient |
| SEXBABY* | Sex of baby |
| SITEDIST | Distance between patient's LSOA and provider site code of treatment |
| SITETRET* | Site code of treatment |
| SPELEND | End of spell indicator |
| SUSCOREHRG | SUS generated Core Spell HRG |
| SUSHRG | SUS generated HRG |
| SUSHRGVERS | SUS generated HRG version number |
| TRETSPEF* | Treatment specialty |
| **LAUNCHES derived field name** | **Description** |
| MATCH_RANK | Quality code of the linkage between the NCHDA record and the HES ID |
| STUDY_ID | LAUNCHES record ID of the linked NCHDA record |
| MY_DOB | Date of Birth - month and year |

| | |
|---|---|
| age_admission | Age at admission to hospital to 4 decimal places (calculated from ADMIDATE and DOB) |
| age_epistart* | Age at episode start to 4 decimal places (calculated from ADMIDATE and DOB) |
| age_discharge | Age at discharge from hospital to 4 decimal places (calculated from DISDATE and DOB) |

Note: The HES/ONS linked data and linkage quality report were received on 17 September 2020. Records with a star (*) were obtained on DD/MM/YYYY.

**Table S3** HES Outpatient (HES OP) fields obtained.

| HES Outpatient field name | Description |
|---|---|
| ATENTYPE | Attendance type |
| ATTENDED | Attended or did not attend |
| ATTENDKEY* | Record identifier |
| DIAG_NN | Diagnosis |
| ENCRYPTED_HESID | Patient identifier - HES generated (encrypted version) |
| ETHNOS* | Ethnic category |
| ETHRAW* | Ethnic character (audit version) |
| FIRSTATT | First attendance |
| HRGNHS | Trust derived HRG value |
| HRGNHSVN | Version No. of Trust derived HRG |
| IMD04 | IMD Index of Multiple Deprivation |
| IMD04_DECILE | IMD Decile Group |
| IMD04RK | IMD Overall Ranking |
| LOCTYPE | Location type |
| MAINSPEF | Main specialty |
| OUTCOME | Outcome of attendance |
| PRIORITY | Priority type |
| PROCODE5 | Provider code (5 character) |
| PROCODET | Provider code of treatment |
| PROTYPE | Provider type |
| PROVDIST | Distance between patient's LSOA and provider |
| REFSOURC | Source of referral |
| RESGOR | Government Office region of residence |
| RESGOR_ONS | Government office region of residence (ONS) |
| RURURB_IND | Rural / urban indicator |
| SEX* | Sex of patient |
| SITEDIST | Distance between patient's LSOA and provider site code of treatment |
| SITETRET* | Site code of treatment |
| SUSHRG | SUS generated HRG |
| SUSHRGVERS | SUS generated HRG version number |
| TRETSPEF | Treatment specialty |
| **LAUNCHES derived field name** | **Description** |
| MATCH_RANK | Quality code of the linkage between the NCHDA record and the HES ID |
| STUDY_ID | LAUNCHES record ID of the linked NCHDA record |
| MY_DOB | Date of Birth - month and year |
| age_appointment_OP | Age at outpatient appointment date to 4 decimal places (calculated from APPTDATE and DOB) |

Note: The HES/ONS linked data and linkage quality report were received on 17 September 2020.
Records with a star (*) were obtained on DD/MM/YYYY.

**Table S4** HES A&E (HES AE) fields obtained.

| HES A&E field name | Description |
|---|---|
| AEARRIVALMODE | Arrival mode |
| AEATTEND_EXC_PLANNED | Attendances excluding planned |
| AEDEPTTYPE | Department type |
| AEKEY | Record identifier |
| DIAG_NN* | A&E diagnosis |
| DIAG2_NN | A&E diagnosis: 2 character |
| DIAGSCHEME* | Diagnosis Scheme in Use |
| DOMPROC | Dominant procedure |
| ENCRYPTED_HESID | Patient identifier – HES generated (encrypted version) |
| EPIKEY | Record identifier |
| ETHRAW* | Ethnic category |
| HRGNHS | Trust derived HRG value |
| HRGNHSVN | Version No. of trust derived HRG |
| IMD04 | IMD Index of Multiple Deprivation |
| IMD04_DECILE | IMD Decile group |
| IMD04RK | IMD Overall rank |
| PROCODE5 | Provider code (5 character) |
| PROCODET | Provider code of treatment |
| PROTYPE | Provider type |
| PROVDIST | Distance between patient's LSOA and provider |
| RESGOR | Government Office region of residence |
| RESGOR_ONS | Government office region of residence (ONS) |
| RURURB_IND | Rural/Urban Indicator |
| SEX* | Sex of patient |
| SITEDIST | Distance between patient's LSOA and provider site code of treatment |
| SITETRET* | Site code of treatment |
| SUSHRG | SUS generated HRG |
| SUSHRGINFO | SUS generated HRG for information |
| SUSHRGVERINFO | SUS generated HRG for information version number |
| SUSHRGVERS | SUS generated HRG version number |
| **LAUNCHES derived field name** | **Description** |
| MATCH_RANK | Quality code of the linkage between the NCHDA record and the HES ID |
| STUDY_ID | LAUNCHES record ID of the linked NCHDA record |
| MY_DOB | Date of Birth - month and year |
| age_arrival_AE | Age at arrival to A&E to 4 decimal places (calculated from ARRIVALDATE and DOB) |

Note: The HES/ONS linked data and linkage quality report were received on 17 September 2020. Records with a star (*) were obtained on DD/MM/YYYY.

**Table S5** ONS mortality fields obtained.

| ONS field name | Description |
|---|---|
| ID | Patient identifier for ONS data – HES generated (encrypted) |
| STUDY_ID | LAUNCHES record ID of the linked NCHDA record |
| ENCRYPTED_HESID | Patient identifier – HES generated (encrypted version) |
| HES_ONS_Match_Rank | Quality code of the linkage between the ONS and HES IDs |
| life_status* | Life status (A=Alive, D=Death) |
| age_life_status | Age at life status to 4 decimal places |
| COMMUNAL_ESTABLISHMENT_NAME | Communal Establishment (with place of death from the communal establishment lookup reference data) |

Note: The HES/ONS linked data and linkage quality report were received on 17 September 2020. Records with a star (*) were obtained on DD/MM/YYYY.

**Table S6** PICANet fields obtained.

| PICANet field name | Description |
|---|---|
| AcuteNec | Acute Necrotising Enterocolitis (NEC) main reason for ICU admission 1 indicates a positive response |
| AdType | Type of admission to PICU |
| bp_PrimReason | Primary reason for admission, a PIM variable used as an input value for PIM2, The PICANet dataset stopped collecting this variable when PIM3 data collection was initiated. This variable is used if present, if not present this value can be assumed as recovering from surgery if the PIM3 variables indicate that the patient was recovering from surgery |
| Cardiomyocarditis | Cardiomyopathy or myocarditis 1 indicates a positive response |
| CareAreaAd | Care area of admission - expected if Source of admission is 1 or 2 |
| ClinicalCodeType | The type of clinical code supplied to PICANet |
| Diagnosis | Read CTV3 clinical code |
| DiagnosticGroup | As Per PICANet annual report dataset reporting categories - only included for primary diagnosis |
| DisPalCare | Discharged for palliative care |
| EcmoDaysDays | Number of days that the patient received Extracorporeal membrane oxygenation (ECMO) |
| ElectiveAd | Elective admission, 1 indicates that the admission was elective |
| Ethnic | Recorded ethnic category of record from |
| FU30DisStatus | Follow up 30 days post discharge |
| FU30Location | Location at 30 days following discharge |
| Gest | Gestational age in weeks, expected between 24 and 42, 99 indicates that the value is unknown |
| HiFlowNasalDays | Number of days that the patient received High flow nasal cannula therapy |
| HypoPlas | Hypoplastic left heart syndrome 1 indicates a positive response |
| IcpDeviceDays | Number of days that the patient used ICP-intracranial pressure monitoring, Intraventricular catheter or external ventricular drain |
| InfInotropeDays | Number of days that the patient received Continuous infusion of inotrope, vasodilator or prostaglandin |
| IntTracheostomy | Tracheostomy performed |
| Intubation | Patient was intubated |
| InvasiveVentilationDay | Number of days that the patient was invasively ventilated based on the PICANet daily interventions fields Invasive ventilation via endotracheal tube, Invasive ventilation via tracheostomy tube, Advanced ventilatory support (jet ventilation) and Advanced ventilatory support (oscillatory ventilation) |
| IsReAd | This record is an emergency readmission |
| LvadDays | Number of days that the patient used Ventricular assist device (VAD) |
| MechVent | Mechanical ventilation |
| MedHistEvid | Is evidence available of past medical history? |
| Mult | Multiplicity of birth, Expecting a value between 1 and 4m 9 indicates that the value is unknown |
| NonInvasiveVentilationDay | Number of days that the patient received Non-invasive ventilatory support |
| PICANetPatientID | Unique identifier for each patient in the PICANet database - pseudonymised from PICANet PatientID |
| PICANetRecordID | Unique identifier for each PICANet Admission Event - pseudonymised from PICANet EventID |
| PICUOrg | PICANet Organisation identifier |
| PICUOrgName | Name of PICANet organisation |
| PIM | Original PIM score |

| | |
|---|---|
| PIM2 | Original PIM2 score |
| PIM2r2011 | PIM2 recalculated 2011 |
| PIM2r2012 | PIM2 recalculated 2012 |
| PIM2r2013 | PIM2 recalculated 2013 |
| PIM2r2014 | PIM2 recalculated 2014 |
| PIM2r2015 | PIM2 recalculated 2015 |
| PIM2r2016 | PIM2 recalculated 2016 |
| PIM2r2017 | PIM2 recalculated 2017 |
| PIM3 | Original PIM3 score |
| PIMr | PIM recalculated |
| PrecedCpr | Cardiac arrest before ICU admission 1 indicates a positive response |
| PreceHospCardArr | Cardiac arrest OUT of hospital 1 indicates a positive response |
| PrevIcuAd | Previous ICU admission |
| RenalSupportDays | Number of days that the patient has received peritoneal dialysis, Haemofiltration, Haemodialysis, Plasma filtration or Plasma exchange |
| Retrieval | Was the patient retrieved to the organisation |
| SpontCerebHaem | Spontaneous cerebral haemorrhage 1 indicates a positive response |
| TotalUnplannedExtubations | The total number of times during an admission that there was dislodgement of the ETT from the trachea, without the intention to extubate immediately and without the presence of airway competent clinical staff appropriately prepared for the procedure occurs |
| TrachDays | Number of Days that the patient had a Tracheostomy cared for by nursing staff |
| UnitDisDest | Destination following discharge |
| UnitDisStatus | Status at discharge |
| UZ01Z_days | number of days the patient received care at UZ01Z |
| XB01Z_days | number of days the patient received care at XB01Z |
| XB02Z_days | number of days the patient received care at XB02Z |
| XB03Z_days | number of days the patient received care at XB03Z |
| XB04Z_days | number of days the patient received care at XB04Z |
| XB05Z_days | number of days the patient received care at XB05Z |
| XB06Z_days | number of days the patient received care at XB06Z |
| XB07Z_days | number of days the patient received care at XB07Z |
| XB09Z_days | number of days the patient received care at XB09Z |
| **LAUNCHES derived field name** | **Description** |
| AgeYrAdmit | Age at admission in years to 4 decimal places |
| AgeYrDeath | Age at death in years to 4 decimal places |
| AgeYrDischarge | Age at discharge in years to 4 decimal places |

Note: The PICANet linked data and linkage quality report were received on 17 October 2019.

**Table S7** ICNARC-CMP fields obtained.

| ICNARC-CMP field name | Description |
|---|---|
| AP2aps | APACHE II acute physiology score |
| AP2probIC3 | APACHE II (ICNARC 2013) probability |
| AP2score | APACHE II score |
| CMPrecordID | CMP (ICNARC) record ID for LAUNCHES |
| IM2018prob | Recalibrated 2018 model predicted mortality probability |
| IMscore | ICNARC model physiology score |
| MEWS | Modified Early Warning Score |
| ODacid_v3 | Metabolic acidosis organ dysfunction |
| ODcardio_v3 | Cardiovascular organ dysfunction |
| ODhaem_v3 | Haematological organ dysfunction |
| ODrenal_v3 | Renal organ dysfunction |
| ODresp_v3 | Respiratory organ dysfunction |
| POPscore | Pancreatitis Outcome Prediction (POP) Score |
| SIRShr_v3 | SIRS tachycardia flag |
| SIRSrr_v3 | SIRS tachypnoea |
| SIRStemp_v3 | SIRS fever or hypothermia flag |
| SIRSwbc_v3 | SIRS white blood cell flag |
| SOFAcardio_v3 | SOFA cardiovascular score (0-2) |
| SOFAcoag_v3 | SOFA coagulation score (0-2) |
| SOFAincr_v3 | SOFA increase from baseline (0-2 per organ) |
| SOFAliver_v3 | SOFA liver score (0-2) |
| SOFAneuro_v3 | SOFA neurological score (0-2) |
| SOFAod_v3 | SOFA number of organ dysfunctions (2+ each organ) |
| SOFArenal_v3 | SOFA renal score (0-2) |
| SOFAresp_v3 | SOFA respiratory score (0-2) |
| SOFAtot_v3 | SOFA total score (0-2 per organ) |
| acsd | Calendar days of advanced cardiovascular support while in your unit |
| admtype | Type of admission |
| ahlos | All hospital length of stay, days |
| ahlosa | Any hospital length of stay after discharge from icu, days |
| ahlosb | Any hospital length of stay before admission to icu, days |
| ahsurv | Ultimate hospital survival |
| arsd | Calendar days of advanced respiratory support while in your unit |
| aulos | All unit length of stay, days |
| ausurv | Ultimate unit survival |
| bcsd | Calendar days of basic cardiovascular support while in your unit |
| bmi | BMI (kg/m2) |
| brsd | Calendar days of basic respiratory support while in your unit |
| bsdtp | Brainstem death declared |
| ccl0d | Days of level 0 care while in your unit |
| ccl1d | Days of level 1 care while in your unit |
| ccl2d | Days of level 2 care while in your unit |
| ccl3d | Days of level 3 care while in your unit |

| | |
|---|---|
| classns | Classification of surgery |
| cpr_v3 | Cardiopulmonary resuscitation within 24 hours prior to admission to your unit |
| crpreg | Admission currently/recently pregnant |
| curb65 | CURB 65 score |
| delay | Length of delay (days) |
| deldis12 | Delayed discharges (12 hour delay) |
| deldis24 | Delayed discharges (24 hour delay) |
| deldis24_exooh | Delayed discharges (24 hour delay), excluding night discharges |
| deldis4 | Delayed discharges (4 hour delay) |
| dep | Dependency prior to admission to acute hospital |
| desc | ICNARC diagnostic category description (raicu1) |
| desth_v3 | Destination following discharge from your hospital |
| dis | Status at discharge from your unit |
| dobest | Date of birth estimated |
| dsd | Calendar days of dermatological support while in your unit |
| ethnic | Ethnicity |
| gsd | Calendar days of gastrointestinal support while in your unit |
| hloca | Hospital housing non-transient location (in) |
| hlocd | Hospital housing non-transient location (out) |
| hrg | Healthcare Resource Group |
| htloca | Hospital housing transient location (in) |
| imd2015 | Quintile of English IMD 2015/Welsh IMD 2014/NI MDM 2010 |
| imd_error | Postcode available for derivation of IMD |
| infection_v3 | Infection |
| itw_v3 | Treatment withheld/withdrawn |
| kdigo_mdrd75 | AKI stage (KDIGO) |
| korgfail | Number of Knaus organ system failures |
| leva | Highest level of care received in the first 24 hours in your unit |
| loca | Location (in) |
| locd | Location (out) |
| lsd | Calendar days of liver support while in your unit |
| nlb | Number of live births (babies) from recent pregnancy |
| npcs | Number of previous Caesarean sections excluding most recent pregnancy |
| nplsb | Number of live births (babies) or stillbirths from previous pregnancies |
| nsb | Number of stillbirths from recent pregnancy |
| nsd | Calendar days of neurological support while in your unit |
| nuaib | Number of unit-acquired infections present in blood |
| orgdys_v3 | Number of organ dysfunctions |
| outrp | Outcome of recent pregnancy |
| ploca | Prior location (in) |
| raicu1 | Primary reason for admission to your unit |
| rdis_v3 | Reason for discharge from your unit |
| readearly | Early readmissions |
| readlate | Late readmissions |
| resa | Residence prior to admission to an acute hospital |
| resd | Residence post-discharge from your hospital |

| | |
|---|---|
| rsd | Calendar days of renal support while in your unit |
| sepsis3_v3 | Sepsis-3 |
| sepsis_v3 | Sepsis |
| sirs_v3 | SIRS criteria count |
| soha | Sector of other hospital (in) |
| sohd | Sector of other hospital (out) |
| sshock3_v3 | Septic shock (Sepsis-3) |
| tnessd | Timeliness of discharge from your unit |
| typeiha | Type of adult ICU/HDU (in) |
| typeihd | Type of adult ICU/HDU (out) |
| version | ICMPDS version number |
| withinsh | Readmission within same hospital stay |
| wkg | Weight (kg) |
| yhlos | Your hospital length of stay, days |
| yhlosa | Your hospital length of stay after discharge from icu, days |
| yhlosb | Your hospital length of stay before admission to icu, days |
| yhsurv | Your hospital survival |
| yulos | Your unit length of stay, hours |
| yusurv | Your unit survival |
| **LAUNCHES derived field name** | **Description** |
| age_ah | Age at admission to hospital (not time-based, 4 decimal places) |
| age_aicu | Age at admission to ICU (time-based, 4 decimal places) |
| age_dbsd | Age at declaration of brainstem death (time-based, 4 decimal places) |
| age_dh | Age at discharge from hospital (not time-based, 4 decimal places) |
| age_dicu | Age at discharge from ICU (time-based, 4 decimal places) |
| age_oah | Age at original admission to hospital (not time-based, 4 decimal places) |
| age_oaicu | Age at original admission to ICU (not time-based, 4 decimal places) |
| age_od | Age at death (time-based, 4 decimal places) |
| age_tw | Age when decision to withdraw treatment made (time-based, 4 decimal places) |
| age_udicu | Age at death (time-based, 4 decimal places) |
| Country | Country |
| Mob | Month of birth |
| qcode_CMP | Quality of CMP record for linkage |
| Yob | Year of birth |

Note: The ICNARC-CMP linked data and linkage quality report were received on 4 December 2019.

**Table S8** NCHDA to PICANet lookup of organisations, used to match hospital patient identifiers.

| NCHDA organisation name | PICANet organisation name |
|---|---|
| ACH. Alder Hey Hospital | PIC010. Liverpool Alder Hey |
| BCH. Birmingham Children’s Hospital | PIC001. Birmingham Children's Hospital |
| BRC. Bristol Children's Hospital | PIC003. Bristol Royal Hospital for Children |
| FRE. Freeman Hospital | PIC021. Newcastle Freeman Hospital |
| GEO. St George's Hospital | PIC015. London St George's Hospital |
| GOS. Great Ormond Street Hospital for Children | PIC011/PIC039. London Great Ormond Street Hospital PICU_NICU/CCCU |
| GRL. Glenfield Hospital | PIC008. Leicester Glenfield Hospital |
| GUY. Evelina London Children's Hospital | PIC012. London Evelina Children’s Hospital |
| HRI. Hull Royal Infirmary | PIC028. Hull Royal Infirmary |
| KCH. King's College Hospital | PIC013. London Kings College Hospital |
| LGI. Leeds General Infirmary | PIC006. Leeds General Infirmary |
| MRI. Manchester Royal Infirmary | PIC018. Manchester Royal Children's Hospital |
| NGS. Northern General Hospital | PIC025 / PIC029. Sheffield General NICU/PICU |
| NHB. Royal Brompton Hospital | PIC014. London Royal Brompton Hospital |
| RAD. John Radcliffe Hospital | PIC024. Oxford John Radcliffe Hospital |
| SBH. St Bartholomew’s Hospital | PIC032. London The Royal London Hospital |
| SGH. Southampton General Hospital | PIC026. Southampton Children’s Hospital |
| STM. St Marys Hospital, Paddington | PIC016. London St Mary's Hospital |
| STO. University Hospital of North Staffordshire | PIC027. Stoke on Trent - Royal Stoke University Hospital |
| UHW. University Hospital of Wales | PIC005. Cardiff Noah’s Ark children’s Hospital for Wales |

**Table S9** HES 8-step linkage method.

| Match_Rank | NHS number | DoB | Sex | Postcode | Extra Condition |
|---|---|---|---|---|---|
| **1** | Exact | Exact | Exact | Exact | |
| **2** | Exact | Exact | Exact | | |
| **3** | Exact | Partial | Exact | Exact | |
| **4** | Exact | Partial | Exact | | |
| **5** | Exact | | | Exact | |
| **6** | | Exact | Exact | Exact | where NHS does not contradict the match and DOB is not 1 January and the POSTCODE is not in the 'ignore' list |
| **7** | | Exact | Exact | Exact | where NHS does not contradict the match and DOB is not 1 January |
| **8** | Exact | | | | |

Note: Sex was not part of the NCHDA identifiers approved for linkage, so only steps 5 and 8 were used.

**Table S10** ICD-10 diagnosis codes indicating congenital heart disease (CHD) or potential mistakenly coded acquired (non-rheumatic) heart disease.

| ICD-10 congenital heart disease code | Description |
|---|---|
| Q20 | Congenital malformations of cardiac chambers and connections |
| Q20.0 | Common arterial trunk |
| Q20.1 | Double outlet right ventricle |
| Q20.2 | Double outlet left ventricle |
| Q20.3 | Discordant ventriculoarterial connection |
| Q20.4 | Double inlet ventricle |
| Q20.5 | Discordant atrioventricular connection |
| Q20.6 | Isomerism of atrial appendages |
| Q20.8 | Other congenital malformations of cardiac chambers and connections |
| Q20.9 | Congenital malformation of cardiac chambers and connections, unspecified |
| Q21 | Congenital malformations of cardiac septa |
| Q21.0 | Ventricular septal defect |
| Q21.1 | Atrial septal defect |
| Q21.2 | Atrioventricular septal defect |
| Q21.3 | Tetralogy of Fallot |
| Q21.4 | Aortopulmonary septal defect |
| Q21.8 | Other congenital malformations of cardiac septa |
| Q21.9 | Congenital malformation of cardiac septum, unspecified |
| Q22 | Congenital malformations of pulmonary and tricuspid valves |
| Q22.0 | Pulmonary valve atresia |
| Q22.1 | Congenital pulmonary valve stenosis |
| Q22.2 | Congenital pulmonary valve insufficiency |
| Q22.3 | Other congenital malformations of pulmonary valve |
| Q22.4 | Congenital tricuspid stenosis |
| Q22.5 | Ebstein anomaly |
| Q22.6 | Hypoplastic right heart syndrome |
| Q22.8 | Other congenital malformations of tricuspid valve |
| Q22.9 | Congenital malformation of tricuspid valve, unspecified |
| Q23 | Congenital malformations of aortic and mitral valves |
| Q23.0 | Congenital stenosis of aortic valve |
| Q23.1 | Congenital insufficiency of aortic valve |
| Q23.2 | Congenital mitral stenosis |
| Q23.3 | Congenital mitral insufficiency |
| Q23.4 | Hypoplastic left heart syndrome |
| Q23.8 | Other congenital malformations of aortic and mitral valves |
| Q23.9 | Congenital malformation of aortic and mitral valves, unspecified |
| Q24 | Other congenital malformations of heart |
| Q24.0 | Dextrocardia |
| Q24.1 | Laevocardia |
| Q24.2 | Cor triatriatum |
| Q24.3 | Pulmonary infundibular stenosis |
| Q24.4 | Congenital subaortic stenosis |

| | |
|---|---|
| Q24.5 | Malformation of coronary vessels |
| Q24.6 | Congenital heart block |
| Q24.8 | Other specified congenital malformations of heart |
| Q24.9 | Congenital malformation of heart, unspecified |
| Q25 | Congenital malformations of great arteries |
| Q25.0 | Patent ductus arteriosus |
| Q25.1 | Coarctation of aorta |
| Q25.2 | Atresia of aorta |
| Q25.3 | Stenosis of aorta |
| Q25.4 | Other congenital malformations of aorta |
| Q25.5 | Atresia of pulmonary artery |
| Q25.6 | Stenosis of pulmonary artery |
| Q25.7 | Other congenital malformations of pulmonary artery |
| Q25.8 | Other congenital malformations of great arteries |
| Q25.9 | Congenital malformation of great arteries, unspecified |
| Q26 | Congenital malformations of great veins |
| Q26.0 | Congenital stenosis of vena cava |
| Q26.1 | Persistent left superior vena cava |
| Q26.2 | Total anomalous pulmonary venous connection |
| Q26.3 | Partial anomalous pulmonary venous connection |
| Q26.4 | Anomalous pulmonary venous connection, unspecified |
| Q26.5 | Anomalous portal venous connection |
| Q26.6 | Portal vein-hepatic artery fistula |
| Q26.8 | Other congenital malformations of great veins |
| Q26.9 | Congenital malformation of great vein, unspecified |
| Q28.8 | Other specified congenital malformations of circulatory system |
| Q28.9 | Congenital malformation of circulatory system, unspecified |
| Q87.4 | Marfan syndrome |
| Q89.3 | Situs inversus |
| **ICD-10 acquired heart disease code** | **Description** |
| I33 | Acute and subacute endocarditis |
| I330 | Acute and subacute infective endocarditis |
| I339 | Acute and subacute endocarditis, unspecified |
| I34 | Nonrheumatic mitral valve disorders |
| I340 | Nonrheumatic mitral (valve) insufficiency |
| I341 | Nonrheumatic mitral (valve) prolapse |
| I342 | Nonrheumatic mitral (valve) stenosis |
| I348 | Other nonrheumatic mitral valve disorders |
| I349 | Nonrheumatic mitral valve disorder, unspecified |
| I35 | Nonrheumatic aortic valve disorders |
| I350 | Nonrheumatic aortic (valve) stenosis |
| I351 | Nonrheumatic aortic (valve) insufficiency |
| I352 | Nonrheumatic aortic (valve) stenosis with insufficiency |
| I358 | Other nonrheumatic aortic valve disorders |
| I359 | Nonrheumatic aortic valve disorder, unspecified |
| I36 | Nonrheumatic tricuspid valve disorders |

| | |
|---|---|
| I360 | Nonrheumatic tricuspid (valve) stenosis |
| I361 | Nonrheumatic tricuspid (valve) insufficiency |
| I362 | Nonrheumatic tricuspid (valve) stenosis with insufficiency |
| I368 | Other nonrheumatic tricuspid valve disorders |
| I369 | Nonrheumatic tricuspid valve disorder, unspecified |
| I37 | Pulmonary valve disorders |
| I370 | Nonrheumatic pulmonary valve stenosis |
| I371 | Nonrheumatic pulmonary valve insufficiency |
| I372 | Nonrheumatic pulmonary valve stenosis with insufficiency |
| I378 | Other nonrheumatic pulmonary valve disorders |
| I379 | Nonrheumatic pulmonary valve disorder, unspecified |
| I38 | Endocarditis, valve unspecified |
| I39 | Endocarditis and heart valve disorders in diseases classified elsewhere |
| I390 | Mitral valve disorders in diseases classified elsewhere |
| I391 | Aortic valve disorders in diseases classified elsewhere |
| I392 | Tricuspid valve disorders in diseases classified elsewhere |
| I393 | Pulmonary valve disorders in diseases classified elsewhere |
| I394 | Multiple valve disorders in diseases classified elsewhere |
| I398 | Endocarditis, valve unspecified, in diseases classified elsewhere |
| I40 | Acute myocarditis |
| I400 | Infective myocarditis |
| I401 | Isolated myocarditis |
| I408 | Other acute myocarditis |
| I409 | Acute myocarditis, unspecified |
| I41 | Myocarditis in diseases classified elsewhere |
| I410 | Myocarditis in bacterial diseases classified elsewhere |
| I411 | Myocarditis in viral diseases classified elsewhere |
| I412 | Myocarditis in other infectious and parasitic diseases classified elsewhere |
| I418 | Myocarditis in other diseases classified elsewhere |
| I42 | Cardiomyopathy |
| I420 | Dilated cardiomyopathy |
| I421 | Obstructive hypertrophic cardiomyopathy |
| I422 | Other hypertrophic cardiomyopathy |
| I423 | Endomyocardial (eosinophilic) disease |
| I424 | Endocardial fibroelastosis |
| I425 | Other restrictive cardiomyopathy |
| I426 | Alcoholic cardiomyopathy |
| I427 | Cardiomyopathy due to drug and external agent |
| I428 | Other cardiomyopathies |
| I429 | Cardiomyopathy, unspecified |
| I43 | Cardiomyopathy in diseases classified elsewhere |
| I430 | Cardiomyopathy in infectious and parasitic diseases classified elsewhere |
| I431 | Cardiomyopathy in metabolic diseases |
| I432 | Cardiomyopathy in nutritional diseases |
| I438 | Cardiomyopathy in other diseases classified elsewhere |
| I44 | Atrioventricular and left bundle-branch block |

| | |
|---|---|
| I440 | Atrioventricular block, first degree |
| I441 | Atrioventricular block, second degree |
| I442 | Atrioventricular block, complete |
| I443 | Other atrioventricular block |
| I444 | Left anterior fascicular block |
| I445 | Left posterior fascicular block |
| I446 | Other fascicular block |
| I447 | Left bundle-branch block, unspecified |
| I45 | Other conduction disorders |
| I450 | Right fascicular block |
| I451 | Other right bundle-branch block |
| I452 | Bifascicular block |
| I453 | Trifascicular block |
| I454 | Nonspecific intraventricular block |
| I455 | Other specified heart block |
| I456 | Pre-excitation syndrome |
| I458 | Other specified conduction disorders |
| I459 | Conduction disorder, unspecified |
| I46 | Cardiac arrest |
| I4.0 | Cardiac arrest with successful resuscitation |
| I461 | Sudden cardiac death, so described |
| I462 | Cardiac arrest due to underlying cardiac condition |
| I468 | Cardiac arrest due to other underlying condition |
| I469 | Cardiac arrest, cause unspecified |
| I47 | Paroxysmal tachycardia |
| I470 | Re-entry ventricular arrhythmia |
| I471 | Supraventricular tachycardia |
| I472 | Ventricular tachycardia |
| I479 | Paroxysmal tachycardia, unspecified |
| I48 | Atrial fibrillation and flutter |
| I480 | Paroxysmal atrial fibrillation |
| I481 | Persistent atrial fibrillation |
| I482 | Chronic atrial fibrillation |
| I483 | Typical atrial flutter |
| I484 | Atypical atrial flutter |
| I489 | Atrial fibrillation and atrial flutter, unspecified |
| I49 | Other cardiac arrhythmias |
| I490 | Ventricular fibrillation and flutter |
| I491 | Atrial premature depolarization |
| I492 | Junctional premature depolarization |
| I493 | Ventricular premature depolarization |
| I494 | Other and unspecified premature depolarization |
| I495 | Sick sinus syndrome |
| I498 | Other specified cardiac arrhythmias |
| I499 | Cardiac arrhythmia, unspecified |
| I50 | Heart failure |

| | |
|---|---|
| I500 | Congestive heart failure |
| I501 | Left ventricular failure |
| I509 | Heart failure, unspecified |

**Table S11** Total number of NCHDA records and number and percentage of those with a valid NHS number, broken down by country of residence derived from postcode (by NICOR).

| Country of residence from patient postcode | Total NCHDA records | Valid NHS number (number of records) | Valid NHS number (percentage of records) |
|---|---|---|---|
| England | 129,952 | 128,351 | 98.8% |
| Wales | 6,514 | 6,457 | 99.1% |
| Crown Dependencies | 531 | 342 | 64.4% |
| Scotland | 681 | 241 | 35.4% |
| Northern Ireland | 1,085 | 80 | 7.4% |
| Overseas | 1,981 | 144 | 7.3% |
| Missing or invalid postcode | 3,118 | 642 | 20.6% |
| Total | 143,862 | 136,257 | 94.7% |

**Table S12** Quality of NHS numbers in each of the datasets (across all records before linkage): yearly total number of records and percentage of records with valid NHS numbers.

| Financial Year | NCHDA *n (% valid)* | PICANet *n (% valid)* | ICNARC-CMP *n (% valid)* | HES Inpatient *n (% valid)* | HES Outpatient *n (% valid)* | HES A&E *n (% valid)* |
|---|---|---|---|---|---|---|
| **1998** | | | | 11,983,893 (74.3%) | | |
| **1999** | | | | 12,196,270 (80.3%) | | |
| **2000** | 6,422 (85.1%) | | | 12,264,676 (83.2%) | | |
| **2001** | 6,170 (92.5%) | | | 12,337,724 (86.5%) | | |
| **2002** | 6,102 (93.7%) | 2,427 (80.2%) | | 12,712,153 (90.5%) | | |
| **2003** | 7,446 (93.3%) | 8,171 (76.9%) | | 13,295,166 (93.6%) | 51,427,003 (93.8%) | |
| **2004** | 6,936 (94.2%) | 8,846 (75.7%) | | 13,706,450 (95.4%) | 54,420,813 (95.6%) | |
| **2005** | 7,720 (94.4%) | 9,699 (75.9%) | | 14,423,506 (95.7%) | 60,608,403 (96.5%) | |
| **2006** | 8,122 (94.9%) | 10,288 (84.4%) | 81,752 (4.5%) | 14,784,581 (96.4%) | 63,217,226 (97.0%) | |
| **2007** | 7,972 (95.4%) | 11,146 (89.9%) | 93,669 (56.7%) | 15,359,062 (96.7%) | 66,649,484 (97.8%) | 12,318,051 (85.4%) |
| **2008** | 8,324 (95.3%) | 11,742 (90.6%) | 101,926 (82.8%) | 16,232,579 (97.1%) | 74,853,493 (98.1%) | 13,794,072 (88.8%) |
| **2009** | 8,726 (96.2%) | 12,646 (91.6%) | 114,519 (90.4%) | 16,806,196 (97.8%) | 84,198,458 (98.2%) | 15,569,736 (90.1%) |
| **2010** | 8,969 (96.4%) | 13,236 (93.1%) | 135,880 (93.3%) | 17,269,882 (98.2%) | 87,998,505 (98.7%) | 16,244,934 (92.0%) |
| **2011** | 9,107 (96.9%) | 13,390 (94.5%) | 156,181 (95.0%) | 17,465,425 (98.5%) | 90,956,844 (98.8%) | 17,619,708 (93.6%) |
| **2012** | 8,994 (95.9%) | 14,307 (94.6%) | 160,737 (95.8%) | 17,715,046 (98.7%) | 94,091,748 (99.0%) | 18,328,896 (95.1%) |
| **2013** | 9,582 (95.3%) | 14,551 (95.1%) | 170,073 (96.1%) | 18,163,101 (98.7%) | 101,844,824 (99.0%) | 18,517,381 (96.0%) |
| **2014** | 9,641 (95.4%) | 15,659 (94.8%) | 191,585 (96.0%) | 18,731,987 (98.8%) | 107,188,423 (99.2%) | 19,556,781 (95.2%) |
| **2015** | 11,472 (94.8%) | 16,898 (94.9%) | 206,383 (96.1%) | 19,239,608 (98.8%) | 113,298,661 (99.2%) | 20,457,805 (95.9%) |
| **2016** | 12,157 (95.9%) | 16,785 (96.6%) | 217,496 (96.4%) | 19,726,907 (98.5%) | 118,578,912 (99.4%) | 20,886,411 (96.9%) |
| **2017** | | | 223,367 (96.7%) | 20,030,870 (98.3%) | 119,378,895 (99.4%) | 21,278,504 (97.1%) |
| **All years** | *143,862 (94.7%)* | *179,791 (90.5%)* | *1,853.568 (88.7%)* | *314,445,082 (93.8%)* | *1,288,711,692 (98.0%)* | *194,572,279 (93.3%)* |

Note: We do not know the quality of identifiers in ONS mortality data, which we obtained linked to HES data.

**Table S13** Number of linked records in each dataset before quality assurance, by estimated financial year

| Financial Year | NCHDA | PICANet | ICNAR C-CMP | HES Inpatient | HES Outpatient | HES A&E | Total |
|---|---|---|---|---|---|---|---|
| **1998** | 0 | 0 | 0 | 16,498 | 0 | 0 | 16,498 |
| **1999** | 0 | 0 | 0 | 19,902 | 0 | 0 | 19,902 |
| **2000** | 6,421 | 15 | 2 | 29,235 | 0 | 0 | 35,673 |
| **2001** | 6,161 | 11 | 1 | 33,390 | 0 | 0 | 39,563 |
| **2002** | 6,137 | 952 | 0 | 37,128 | 0 | 0 | 44,217 |
| **2003** | 7,402 | 3,226 | 0 | 43,106 | 132,710 | 0 | 186,444 |
| **2004** | 6,968 | 3,464 | 0 | 45,657 | 150,065 | 0 | 206,154 |
| **2005** | 7,684 | 3,828 | 0 | 50,436 | 177,020 | 0 | 238,968 |
| **2006** | 8,152 | 4,052 | 6 | 52,256 | 196,331 | 0 | 260,797 |
| **2007** | 7,984 | 4,136 | 154 | 56,918 | 224,342 | 23,352 | 316,886 |
| **2008** | 8,294 | 4,275 | 215 | 60,251 | 255,702 | 27,597 | 356,334 |
| **2009** | 8,719 | 4,748 | 273 | 65,634 | 294,368 | 32,876 | 406,618 |
| **2010** | 8,987 | 4,891 | 388 | 69,485 | 323,853 | 35,989 | 443,593 |
| **2011** | 9,102 | 5,103 | 407 | 70,851 | 348,802 | 38,968 | 473,233 |
| **2012** | 9,013 | 5,176 | 411 | 71,279 | 370,113 | 41,745 | 497,737 |
| **2013** | 9,593 | 5,435 | 473 | 72,077 | 408,561 | 42,959 | 539,098 |
| **2014** | 9,639 | 5,435 | 447 | 73,027 | 442,349 | 45,018 | 575,915 |
| **2015** | 11,492 | 5,546 | 629 | 76,245 | 470,121 | 47,341 | 611,374 |
| **2016** | 12,114 | 5,504 | 686 | 73,112 | 478,591 | 47,060 | 617,067 |
| **2017** | 0 | 0 | 572 | 52,074 | 426,352 | 43,604 | 522,602 |
| **All years** | *143,862* | *65,797* | *4,664* | *1,068,561* | *4,699,280* | *426,509* | *6,408,673* |

Note: financial years (running from April to March) were estimated using the ages at events and the estimated date of birth (we took day 15$^{th}$ of the known month of birth as date of birth).
